# Identifying the determinants of health protective behaviors during the COVID-19 pandemic using machine learning: an analysis of six countries

**DOI:** 10.64898/2026.05.05.26352439

**Authors:** Joshua M Chevalier, Leonard Stellbrink, Lisanne Steijvers, Senne Wijnen, Florian van Daalen, Lilian Kojan, Nannan Li, Beate Jahn, Uwe Siebert, André Calero Valdez, Mickaël Hiligsmann, Rik Crutzen, Nicole Dukers-Muijrers, Mirjam Kretzschmar

## Abstract

Individuals adapt their behavior in response to infectious disease epidemics. Understanding the determinants of behavior, particularly the impact of infections themselves, can help model the feedback loop between disease and behavior in epidemic models. We combined the Imperial College London YouGov COVID-19 behavior survey with hospitalization data and the Oxford COVID-19 government response tracker stringency index to identify the key predictors of three health behaviors—social distancing, masking, and personal protective measures (e.g. handwashing)— during an early phase of the COVID-19 pandemic in six different countries. We compared two machine learning algorithms—logistic regression with stepwise Akaike Information Criterion and extreme gradient boosting (XGBoost). Top predictors of health behavior were perceived disease severity, hospitalizations, willingness to isolate, and intervention effectiveness, across the six countries. Logistic regression and XGBoost had comparable performance. Machine learning algorithms trained on real-world data could be used to predict individual behavior uptake in agent-based network models.

## Introduction

SARS-CoV-2 emerged in late 2019 and disseminated around the world by early 2020—causing the COVID-19 pandemic. To combat the spread of infection, before vaccines were available, countries implemented non-pharmaceutical interventions (NPIs). These included mandates for mask wearing, gathering restrictions, or the closure of schools and other public spaces (1). Authorities also promoted voluntary health behaviors—hand washing, covering coughs/sneezes, reducing social contacts, and staying home (1). Adherence and compliance to mandatory and voluntary NPIs have been widely studied to understand the determinants of health behavior participation.

Understanding what drives human behavior can help us to more effectively implement and target NPIs during future pandemics.

Psychosocial theories that attempt to explain health behaviors include the health belief model (HBM), the theory of planned behavior (TPB), and protection motivation theory (PMT) (2–4). These theories categorize people’s motivations into broad categories—perceived threat, social norms, attitudes, self-efficacy, barriers, or associated costs and benefits. Prior studies on the determinants of health behavior identified features that fall within these motivations. Bish and Michie conducted a review on pandemic related health behaviors and found that demographics afected health behavior—age, sex, ethnicity, and education—but also that perceived threat (susceptibility, severity), belief in the effectiveness of the intervention, and trust in authorities were associated with behavior (5). Hanratty et al. found associations between sex, perceived threat, and self-efficacy, and hand-washing, face coverings, and social distancing in the context of pandemic influenza (6). Further, they concluded these determinants largely aligned with the Health Belief Model and Protection Motivation Theory. In a more recent review, Hanratty et al. found relationships between attitudes, social norms, perceived behavioral control, response effectiveness, and self-efficacy, and distancing behaviors (7); while a similar review on masking behavior identified relationships with perceived benefits and effectiveness of the behavior (8). Cabrera-Álvarez et al. found social norms, trust in science, perceived intervention effectiveness, and perceived risk were all significant predictors of social distancing behavior during COVID-19 in Spain (9).

The COVID-19 pandemic brought to the forefront mathematical modelling, an influential tool for health policy and decision making, to forecast epidemic transmission and evaluate the impact of interventions. Central to epidemic spread is human behavior, as individuals adapt their behavior to avoid infection which ultimately influences the disease dynamics. Therefore, a feedback loop exists between the spread of a disease and the uptake of health behavior. Capturing this feedback loop is important for the formulation of more realistic and accurate mathematical models. Theoretical frameworks have been proposed to mechanistically represent these feedback loops (10–14). Perra et al. proposed compartmental frameworks in which the fear of disease spreads locally or globally, based on prevalence, wherein individuals transition to behavior practicing compartments after being exposed to fear (12). Funk et al. proposed frameworks in which awareness of the disease spreads amongst individuals and influences the uptake of health behaviors (10,11). Ryan et al. proposed a compartmental model that utilized the health belief model to parameterize the transitions between behavioral compartments (13). While these theoretical frameworks successfully incorporate human behavior into epidemic models, there is little evidence on how to parameterize the behavioral components of these models to accurately reflect behavioral adaptation.

Understanding the determinants of health behavior uptake during the pandemic can aid in parameterizing the feedback loop between disease and behavior. In this analysis we combine two different data sources—a COVID-19 behavioral survey and country-level epidemiological data from the COVID-19 pandemic—to identify the personal characteristics, perceptions, and broader societal epidemic metrics associated with participation in three health behaviors (social distancing, mask wearing, and personal protective measures) across six different countries using two machine learning algorithms. We then consider the potential use of these algorithms to predict behavior uptake among individuals within agent-based network models.

## Methods

### Behavioral Data

The Institute of Global Health and Innovation (IGHI) through Imperial College London partnered with YouGov to collect survey responses on behavior during the COVID-19 pandemic from April 2020 to March 2022 (15). Survey data from 29 countries is publicly accessible on the Imperial College London YouGov Covid-19 Behavior Tracker Data Hub (16). Survey questions focused on demographics, social network composition, individual perceptions, and health behaviors—testing, isolation, social distancing, masking, personal hygiene, and vaccination.

We obtained the secondary data from six different countries—Canada, France, Japan, the Netherlands, Sweden, and the UK. Countries were chosen to reflect various regions of the world but were restricted to those with hospitalization data available from Our World in Data for the dates of the analysis. We filtered the data to focus on the first year of the pandemic, during the second wave, before vaccination became widely available, from 24 June 2020 to 13 January 2021.

Vaccination was excluded because it may have reduced individuals perceived susceptibility to and severity of COVID-19, as well as the continued need for NPIs.

### COVID-19 Data

We extracted the number of COVID-19 hospital patients for Canada, France, Japan, Sweden, and the UK from Our World in Data (17). For the Netherlands, Our World in Data hospitalization data was only available from 2021 so we instead obtained the number of daily COVID-19 hospital admissions from the National Institute of Public Health and the Environment (RIVM) and extrapolated to the daily number of patients in the hospital using a six-day length of stay (18). A six-day length of stay was determined by matching the projected total hospital occupancy with data available from Our World in Data starting from 2021. The Oxford Coronavirus Government Response Tracker (OxCGRT) Stringency Index was also extracted from Our World in Data for all six countries (19). The OxCGRT Stringency Index is a policy indicator of a given country’s response to the COVID-19 pandemic based on 19 policies—closure and containment, health and economic policies—with a value between 0 and 100 (19). COVID-19 hospitalizations and the national stringency index were linked to the behavioral survey data by date for each country. COVID-19 hospitalizations were converted to country relative measures (per 100,000 population).

### Health Protective Behaviors

We sought to understand the determinants of three health protective behaviors elicited in the survey responses—social distancing, wearing a mask or facial covering in public, and personal protective measures (PPM)—practiced over the past seven days. We created a composite score to represent social distancing behavior based on four different questions in the survey: avoided small, medium, and large social gatherings, or crowds. Questions were answered on a five-point Likert scale of agreement, and summed, for a minimum composite score of four or a maximum score of 20. The social distancing variable was converted to binary with a score of 16 or higher equating to practicing social distancing, representing an average answer of “frequently” across the four questions. The masking outcome was expressed in one question: how often one has worn a face mask outside their home, on a five-point Likert scale, converted to binary with a score of four or higher equating to regular masking. PPM was also represented by four questions in the survey: in the past seven days, how often one has washed their hands with soap and water, used hand sanitizer, covered coughs and sneezes, and avoided symptomatic individuals, asked on a five-point Likert scale. Converted to a binary outcome, a composite score of 18 or higher was considered practicing increased PPM, as the average PPM score across all countries in the first week of the survey (April 2020) was 16.9.

### Predictor Variables

Predictor variables—features—were selected from the data based on prior knowledge, expert opinion, those commonly identified in the academic literature, and those related to the themes in psychosocial theories (HBM, TPB, PMT). Variable inclusion was further dependent on the number of response weeks for the related questions to maximize the sample size over time. **Table 1** lists the predictor variables used in the analysis, their respective category, descriptions, and source.

**Table 1.** Description of predictor variables used in machine learning analyses.

| Variable | Description | Category | Source |
| --- | --- | --- | --- |
| Age | Survey participant's age | Demographics | Imperial College<br>London YouGov<br>Covid-19 |
| Gender | Survey participant's gender |  |  |
| Working outside home | Not working from home, unemployed, or retired |  |  |
| Household size | Number of people living in household (1–8+) | Social network | Behaviour Tracker Survey |
| Number children | Number of children living in household (1–6+) |  |  |
| Outside contacts | How many people outside their household they have had physical contact with in past seven days (0–1000) |  |  |
| Physical contact household | How many members of their household they have had physical contact with in past seven days (0–1000) |  |  |
| Preexisting condition | Presence of at least one of 13 pre-existing health conditions | Current health |  |
| Symptoms | COVID-19 like symptoms in past seven days |  |  |
| Health consciousness | Belief practicing behaviors for health is important | Health practices |  |
| Tested covid self | Participant has tested for COVID-19 in past seven days |  |  |
| Tested covid household | Household member has tested for COVID-19 in past seven days |  |  |
| Willing to isolate | Willingness to isolate for seven days if required to do so |  |  |
| Mask effectiveness self | Perceived efficacy of masks to protect oneself from COVID-19 | Perceived intervention effectiveness |  |
| Mask effectiveness others | Perceived efficacy of masks to protect others from COVID-19 |  |  |
| Mask barrier | Inability to wear a mask | Intervention barriers |  |
| Perceived susceptibility | Perceived as likely to contract COVID-19 | Perceived threat |  |
| Perceived severity | Perceived COVID-19 as dangerous |  |  |
| Well-being | Cantril ladder—well-being, life satisfaction | Personal well-being |  |
| Anxiety and depression score | Compositive score of PHQ-4 questions |  |  |
| Government handling | Trust in government handling of pandemic | Government trust |  |
| Government confidence | Confidence in government response to pandemic |  |  |
| Hospital patients per 100,000 population | Number of COVID-19 hospital patients per 100,000 population | COVID-19 pandemic metrics | Our World in Data |
| Stringency index | OxCGRT COVID-19 stringency index (0–100) |  |  |

Variables with more than two levels were included as continuous numeric variables, representing continuous responses (e.g. number of contacts) or agreement on five- or seven-point Likert scales.

### Statistical Analyses

Two machine learning models were used to identify the key statistical predictors of masking, social distancing, and PPM behavior during the COVID-19 pandemic. The predictors identified by these algorithms cannot be confirmed as causal. The data was subset to include all predictor and outcome variables, filtered by the date range, with missing responses omitted. The data was split 80% for training, and 20% for testing. All analyses were performed using R (version 4.4.2).

First, we performed logistic regression using stepwise Akaike Information Criterion with both forward and backward selection for variable inclusion. The standardized odds ratios—where one unit change is equal to one standard deviation in the predictor variable—and their confidence intervals were calculated for each feature. Variance inflation factors (VIF) were calculated for each feature to identify multicollinearity between predictors. P-values and area under the curve (AUC) statistics were extracted. The logistic regression models were tested for their predictive performance on the test data. A predicted probability greater than 0.5 was considered a positive outcome. The probability of the models to successfully predict those who did or did not practice the health behavior (accuracy) was calculated.

We then used extreme gradient boosting (XGBoost), a machine learning algorithm that builds an ensemble of decision trees using gradient boosting, to further investigate the effect of predictor features without the linear assumption of logistic regression. This algorithm was then compared to the standard approach of logistic regression. XGBoost Hyperparameters were optimized for each health behavior model using parallel Bayesian optimization with four-fold cross validation on a pooled subset of observations randomly selected from each country (N=2000 per country). Scoring criteria for hyperparameter selection was minimum log loss. Once optimal hyperparameters were identified, the optimal number of boosting rounds was determined through five-fold cross validation with 1500 boosting rounds and 50 early stopping rounds on the training dataset. If there was no improvement in log loss on the validation fold after 50 rounds, the model would stop and select the last best iteration. The final XGBoost models then utilized the optimal number of boosting rounds on the entire training dataset.

XGBoost models were assessed using SHapely Additive exPlanations (SHAP values) calculated per feature per observation in the training data. Derived from game theory, a SHAP value for a particular feature represents its contribution to the model prediction for a given observation, and can be positive or negative (20). In the case of binary classification, the SHAP values represent the change in log odds. Added together, the feature SHAP values for a given observation represent the log odds of the model’s overall prediction for that observation. SHAP values were used to investigate feature influence on predictive outcomes as well as interactions between features. SHAP interaction values decompose the standard SHAP values to show the contribution of the interaction between two features on the prediction. The average SHAP value for a feature across all observations represents the average directional influence of that feature—positive or negative contribution to the outcome. The average of the absolute SHAP values for a feature across all observations shows the magnitude of impact (overall feature importance on outcome). The average SHAP value per feature, the average of the absolute SHAP values per feature, and the AUC of the overall model, were calculated. The XGBoost models were tested for their predictive performance on the test data.

## Results

Each country had a variable number of survey respondents (4,345–15,283), representing 12–17 survey weeks during the study period, after missing observations were removed (**Appendix Table S1**). Age distribution was similar between countries (mean: 48–50 years, standard deviation: ±15– 17 years), while the gender distribution differed with >50% of female respondents in all countries except Japan (48%; **Appendix Table S1**). Over the course of the study period the national stringency index and the number of COVID-19 hospital patients per 100,000 population generally increased across all countries, illustrating the increase in wildtype COVID-19 infections in the second half of 2020 and the start of the Alpha variant (B.1.1.7) wave in December 2020 (**Appendix Figures S1–S4**). Similarly, the average masking score, social distancing score, and PPM score per survey week increased in all countries during the study period. The binary distributions of the three health behaviors differed across countries (**Appendix Figures S5**). An inverse linear relationship was observed between the social distancing score and self-reported contacts from outside the home (**Appendix Figure S6**).

### Social Distancing Behavior

#### Logistic Regression

Logistic regression analysis with stepwise AIC identified multicollinearity between COVID-19 hospital patients and the national stringency index in the Netherlands (VIF>3), Canada (VIF>4), Japan (VIF>5), and Sweden (VIF>10). Thus, we removed the national stringency index from all logistic regression analysis—including masking and PPM. Across all countries the top three predictors of social distancing behavior were perceived severity, the number of COVID-19 hospital patients, and willingness to isolate (**Figure 1**). In Canada, Japan, Sweden, and the UK the top predictor was perceived severity, while the number of COVID-19 hospital patients was the top predictor in France and the Netherlands. Age was also a significant predictor, with a positive influence on social distancing, in all countries except Japan. Working outside the home was a significant and negative predictor of social distancing across all countries. When the stringency index was included, the standardized odds ratio of COVID-19 hospital patients was inflated. When hospitalizations were removed the stringency index replaced hospitalizations as a top predictor of social distancing behavior (**Appendix Figure S7**).

**Figure 1.**
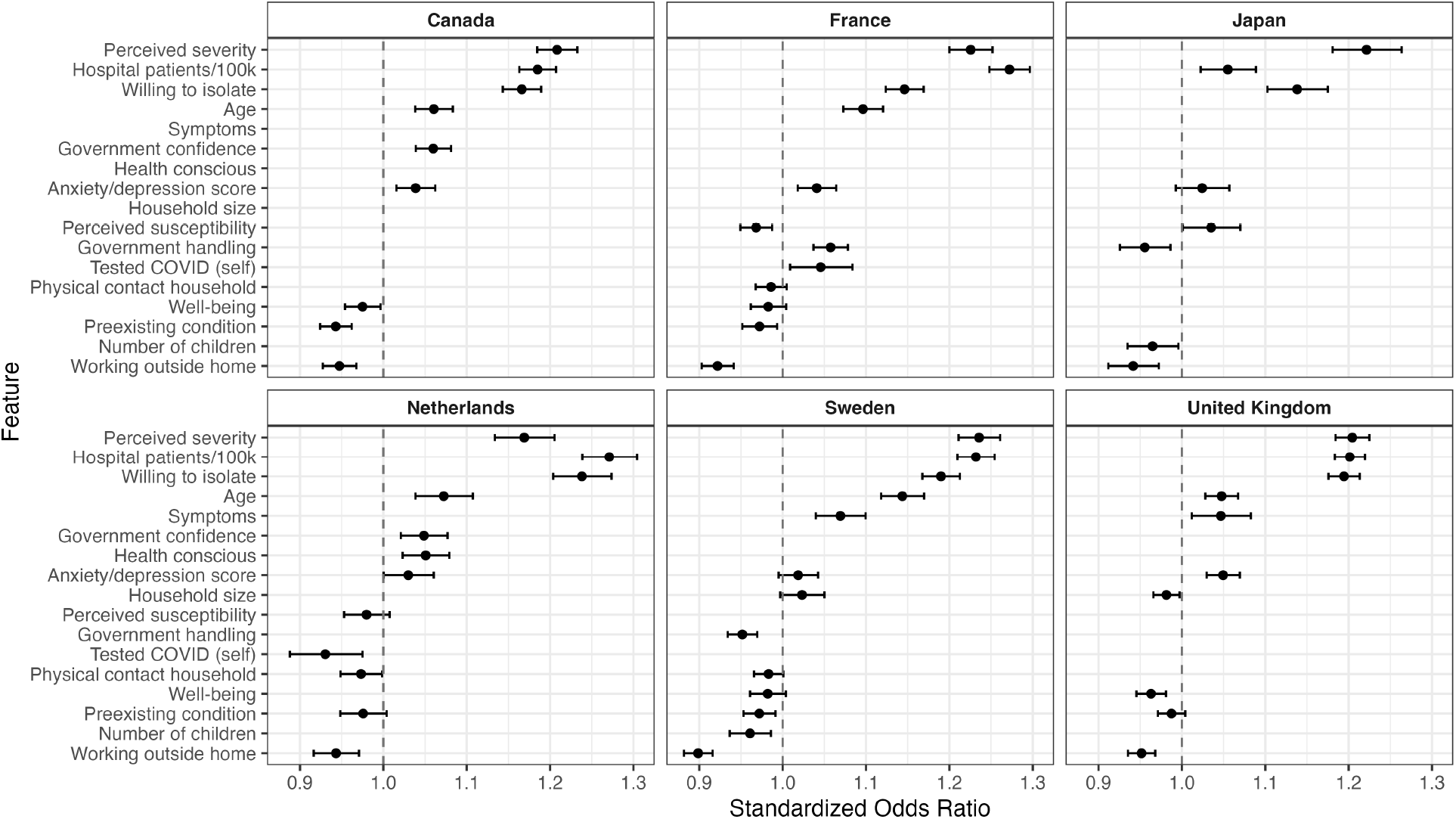
Standardized odds ratios and 95% confidence intervals of features that were included in the final logistic regression model with stepwise AIC for social distancing behavior, by country.

#### XGBoost

XGBoost analysis identified perceived severity, COVID-19 hospital patients, and willingness to isolate as the top three predictors of social distancing in all countries except Japan (**Figure 2**). The national stringency index was included, as XGBoost can handle multicollinearity, but it was not identified as a top predictor. In Japan the top three predictors of social distancing were perceived severity, willingness to isolate, and gender, with COVID-19 hospital patients being the fourth most influential predictor. In all other countries age and working outside the home were also influential predictors. The effect of these features on social distancing behavior (positive/negative contribution) was dependent on the feature values (**Figure 3**). When COVID-19 hospital patients, willingness to isolate, perceived severity, and age increased, the corresponding SHAP values were positive, and when the feature values were low the SHAP values were negative (**Figure 4**). This indicates people may have responded to increasing infection levels and perceived disease severity by social distancing, and those that were already willing to participate in intervention measures (self-isolation) were more likely to social distance.

**Figure 2.**
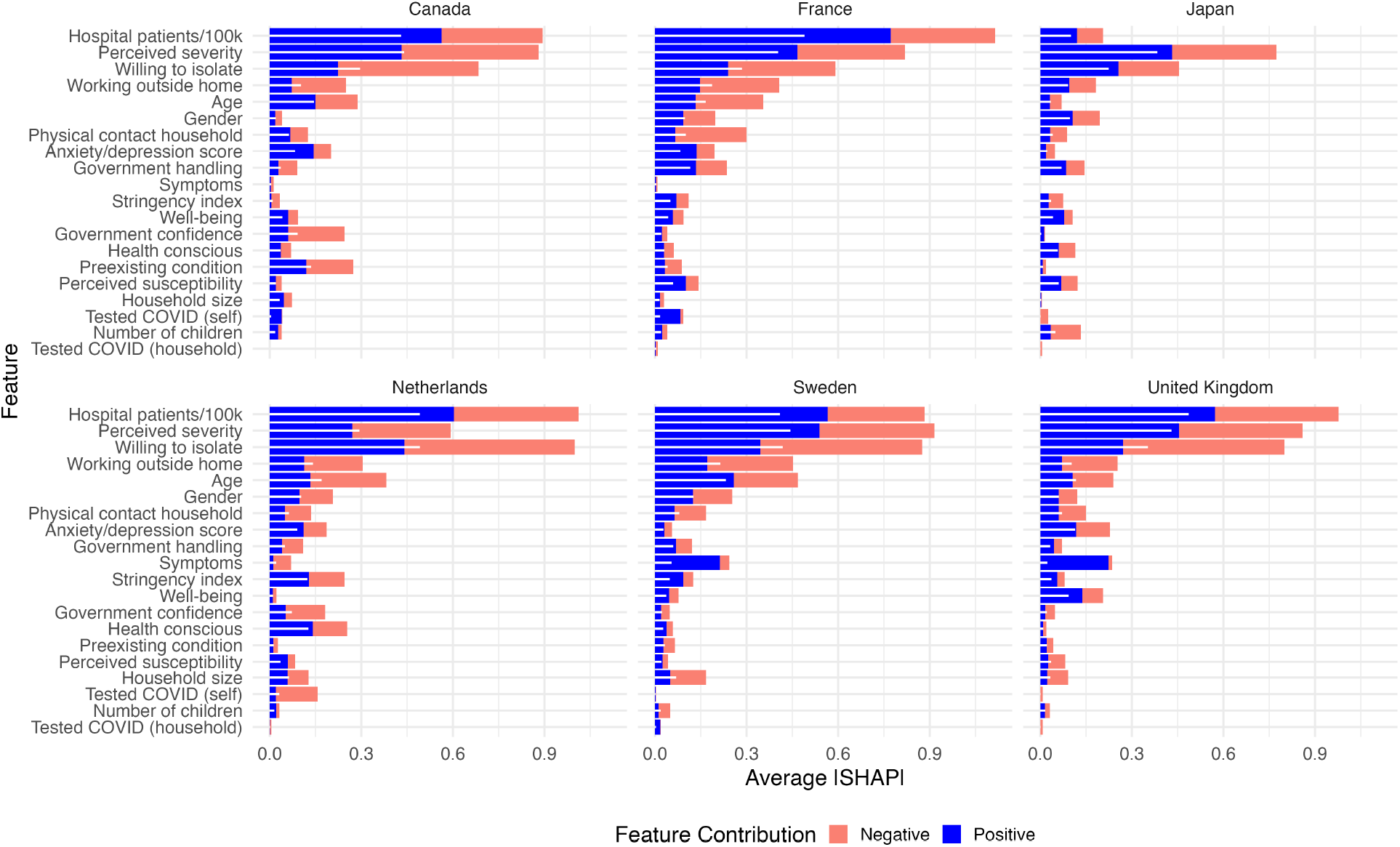
Respective averages of the absolute SHAP values by feature, feature contribution, and country from XGBoost for social distancing. Colors represent feature contribution (positive SHAP: blue, negative SHAP: orange) while white bars show the combined average of the absolute SHAP values for the feature.

**Figure 3.**
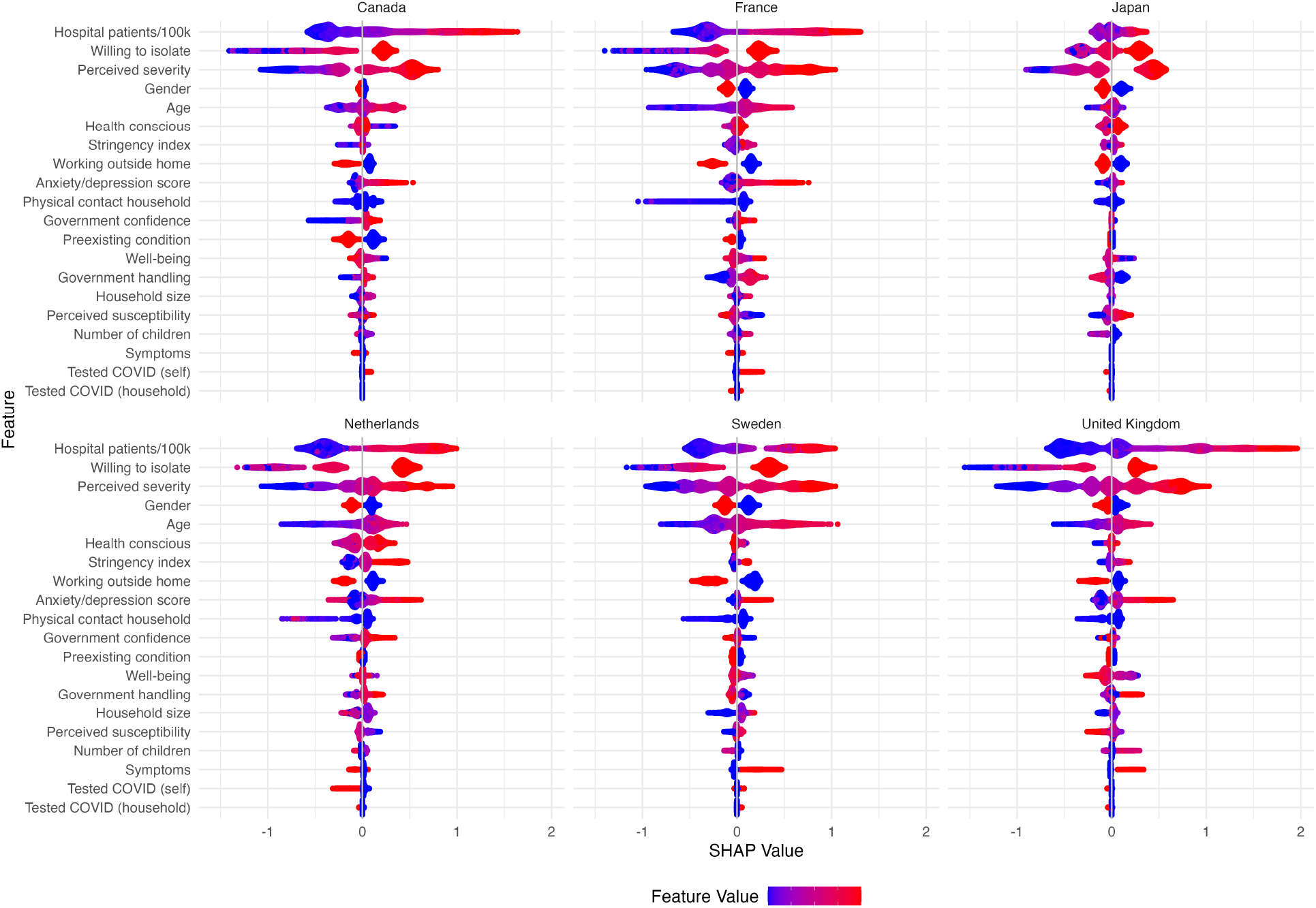
SHAP value distributions for social distancing from XGBoost by observation and feature, colored by feature value across countries.

**Figure 4.**
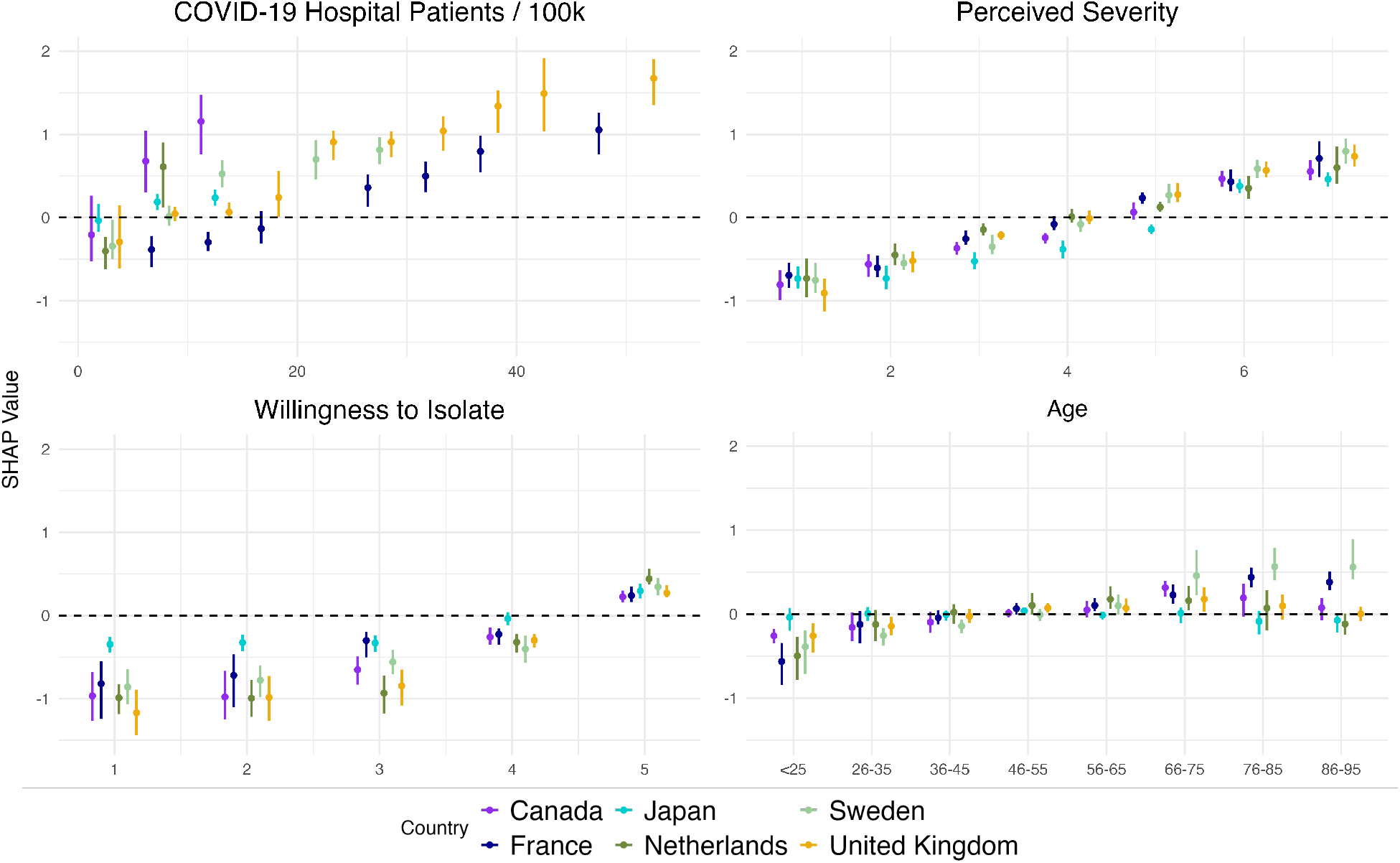
Average SHAP values and 95% quantile range for COVID-19 hospital patients per 100,000, perceived severity, willingness to isolate, and age, by increasing feature value and country (color).

Interactions between predictors were assessed through SHAP interaction values (**Appendix Figure S8**). SHAP interaction values, and further exploration through SHAP dependency plots, revealed no significant interactions between key features.

### Masking behavior

#### Logistic Regression

The top predictor of masking behavior was perceived mask effectiveness at protecting others in Canada, France, and the UK, but perceived mask effectiveness at protecting oneself in Sweden (**Appendix Figure S9**). In the Netherlands the top predictor was the number of COVID-19 hospital patients, while in Japan it was willingness to isolate. Key predictors included mask effectiveness, hospitalizations, perceived severity and willingness to isolate. Inability to wear to mask (mask barrier) was a significant negative predictor across all countries.

#### XGBoost

XGBoost identified COVID-19 hospital patients, perceived mask effectiveness for oneself, and perceived mask effectiveness for others in Canada, Sweden, and the UK (**Figure 5**) as the top predictors of masking behavior. France and the Netherlands were similar with COVID-19 hospital patients, and the national stringency index, but with perceived mask effectiveness for oneself in the Netherlands, and inability to wear a mask (mask barrier) in France. The top three predictors in Japan were willingness to isolate, perceived severity, and the number of contacts from outside the home.

**Figure 5.**
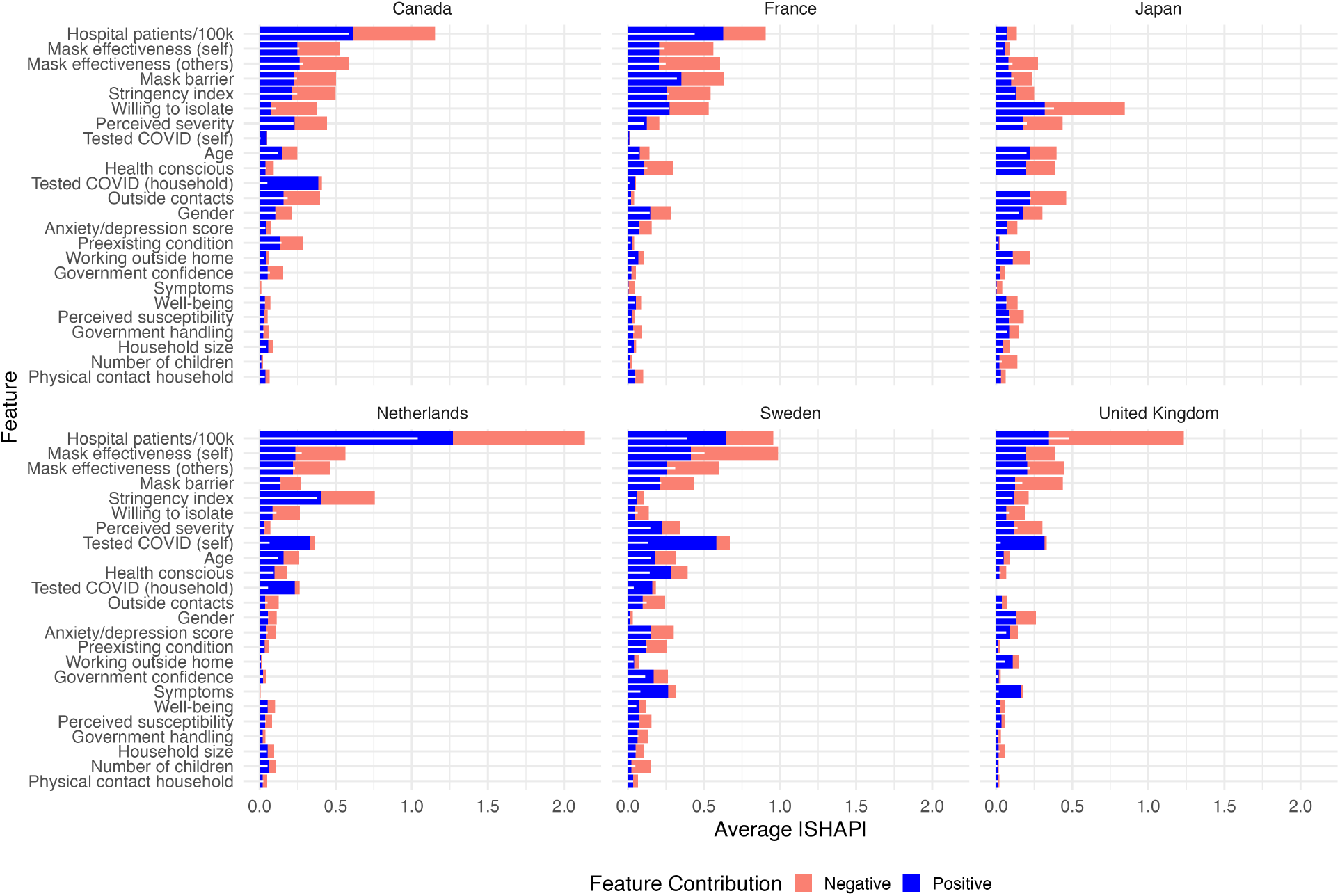
Respective averages of the absolute SHAP values by feature, feature contribution, and country from XGBoost for masking. Colors represent feature contribution (positive SHAP: blue, negative SHAP: orange) while white bars show the combined average of the absolute SHAP values for the feature.

### Personal Protective Measure Behavior

#### Logistic Regression

The top predictor of increased PPM behavior (hand washing/sanitizer, covered coughs/sneezes, avoided symptomatic individuals) as identified through logistic regression was willingness to isolate across all countries, except in France where it was perceived severity (**Appendix Figure S10**). The top four predictors in all countries included willingness to isolate, COVID-19 hospital patients, perceived severity, and health consciousness, all having a positive effect.

#### XGBoost

XGBoost also identified willingness to isolate as the top predictor of increased PPM behavior in all countries, including France (**Figure 6**). Other top predictors of increased PPM included perceived severity, gender, health consciousness, and COVID-19 hospital patients, but the relative importance of these features varied by country.

**Figure 6.**
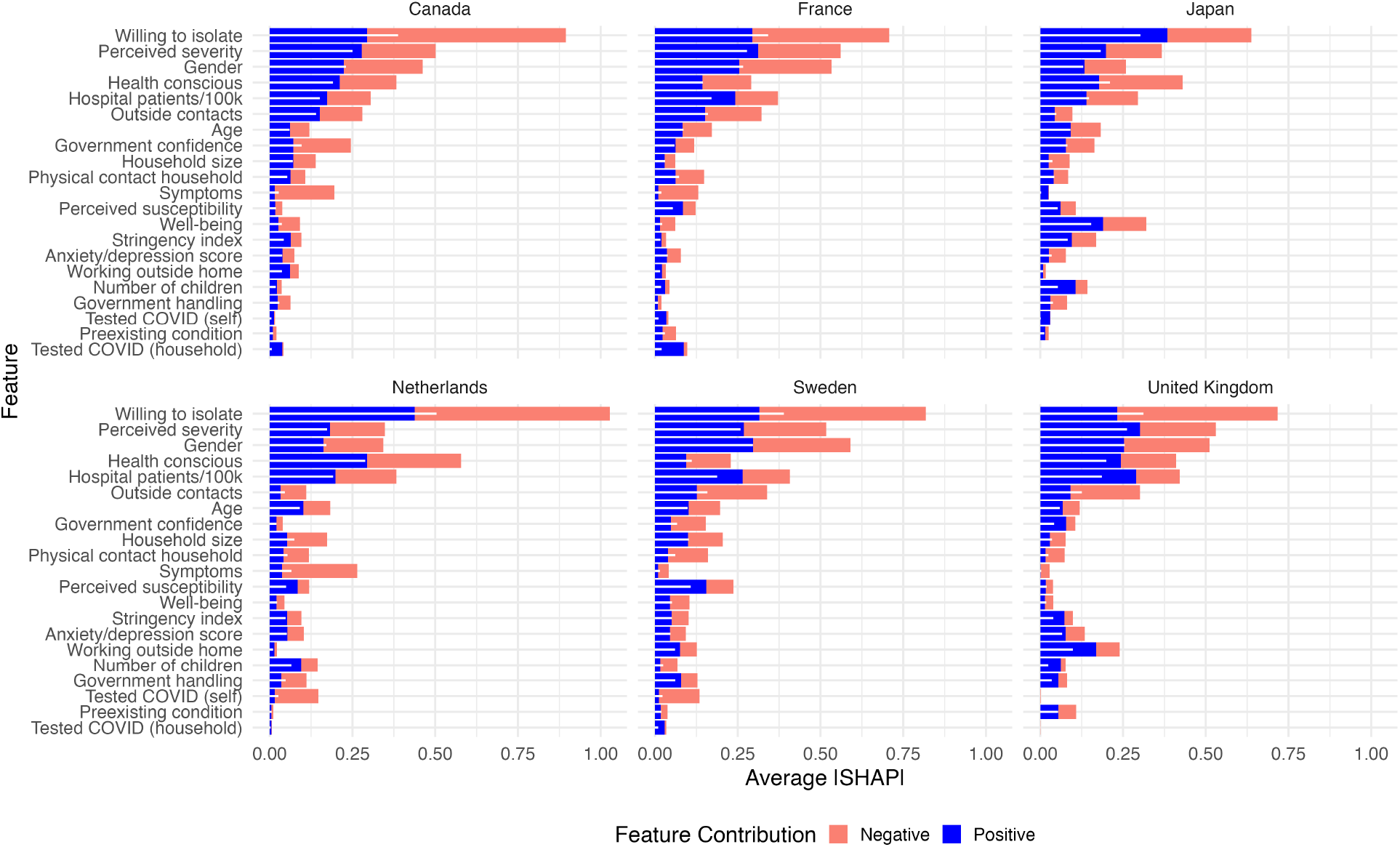
Respective averages of the absolute SHAP values by feature, feature contribution, and country from XGBoost for PPM. Colors represent feature contribution (positive SHAP: blue, negative SHAP: orange) while white bars show the combined average of the absolute SHAP values for the feature.

### Comparison of Machine Learning models

For the three health behaviors, both logistic regression with stepwise AIC and XGBoost identified the same top predictors. With respect to model performance, the AUC, and the predictive accuracy of the models on the test data, logistic regression and XGBoost were comparable with XGBoost having a slight edge across most models (**Table 2**). Models for masking behavior had the highest performance with >75% accuracy across countries, while models for increased PPM behavior had the lowest performance with <70% accuracy in some countries.

**Table 2.** Performance metrics of logistic regression and XGBoost models by country and health behavior.

| Country | Health Behavior | AUC |  | Predictive Probability |  |
| --- | --- | --- | --- | --- | --- |
|  |  | Logistic Regression | XGBoost | Logistic Regression | XGBoost |
| Canada | Masking | 0.768 | 0.837 | 0.846 | 0.873 |
|  | PPM | 0.719 | 0.726 | 0.776 | 0.789 |
|  | Social Distancing | 0.741 | 0.726 | 0.714 | 0.728 |
| France | Masking | 0.788 | 0.818 | 0.906 | 0.910 |
|  | PPM | 0.701 | 0.717 | 0.687 | 0.705 |
|  | Social Distancing | 0.751 | 0.753 | 0.701 | 0.701 |
| Japan | Masking | 0.728 | 0.734 | 0.908 | 0.910 |
|  | PPM | 0.689 | 0.668 | 0.648 | 0.656 |
|  | Social Distancing | 0.686 | 0.674 | 0.648 | 0.649 |
| Netherlands | Masking | 0.859 | 0.890 | 0.803 | 0.817 |
|  | PPM | 0.732 | 0.766 | 0.679 | 0.699 |
|  | Social Distancing | 0.774 | 0.791 | 0.720 | 0.727 |
| Sweden | Masking | 0.836 | 0.852 | 0.902 | 0.904 |
|  | PPM | 0.714 | 0.738 | 0.673 | 0.683 |
|  | Social Distancing | 0.775 | 0.772 | 0.701 | 0.703 |
| United Kingdom | Masking | 0.701 | 0.776 | 0.741 | 0.786 |
|  | PPM | 0.702 | 0.700 | 0.689 | 0.690 |
|  | Social Distancing | 0.754 | 0.762 | 0.732 | 0.739 |

## Discussion

The determinants of health protective behaviors during the COVID-19 pandemic identified in this analysis align with those reported in the prior literature, such as perceived disease severity and intervention effectiveness but also indicate that people respond strongly to a growing epidemic— represented here by the number of COVID-19 hospitalizations (5–8,21). Further, our results suggest some individuals are predisposed to take up health protective behaviors when prompted to do so, represented by their reported willingness to isolate. Epidemiological metrics like case numbers, hospitalizations, and deaths are not frequently included in behavioral determinant studies, despite a known feedback loop (22). The detailed data obtained during the COVID-19 pandemic has given us the opportunity to better explore the relationship between epidemic metrics and health behavior, with the potential to quantify this dynamic for use in transmission models.

COVID-19 hospital patients per 100,000 population was a significant and strong predictor of all health protective behaviors. Together with perceived severity, another strong predictor, they represent the perceived threat an individual has towards infection. This study provides evidence that individuals may respond to increasing infection levels by adopting stricter health protective behaviors. At the same time, country level responses to the pandemic were often based on metrics like hospital and intensive care unit bed occupancy, so the level of hospitalizations was directly related to the national stringency index. VIF-testing identified multicollinearity between COVID-19 hospital patients and the national stringency index. The national stringency index may have further prompted the adoption of health protective behaviors through masking requirements and gathering restrictions, although it is not possible to disentangle the relative effects in this analysis. However, the machine learning algorithm, XGBoost, identified a stronger relationship between hospitalizations and health protective behaviors despite the presence of the stringency index.

An individual’s reported willingness to comply with isolation if required was also a significant and strong predictor of all health behaviors, with an even stronger effect than hospitalizations in some settings. This suggests that some individuals are predisposed to heed authoritative direction and participate in health behaviors when prompted to do so by other externalities. These individuals may also have a greater level of health consciousness, a top predictor in the PPM analysis, which identified individuals who thought partaking in behaviors important to their health (health consciousness) practiced greater PPM. Future analyses, such as latent profile analyses, could identify health conscious or health neutral individuals based on related variables to understand if the rate at which they respond to other external factors—like hospitalizations—differs. Prior mechanistic models have explored such opinion dynamics between health positive and health neutral individuals (23).

The features that influenced the uptake of health protective behaviors were consistent across country contexts, apart from Japan. This was most notable for masking behavior, as masking in public while sick was already a widely practiced behavior in Japan with no additional mandates put in place during the COVID-19 pandemic (24–27). During the survey period, >80% of respondents from Japan reported wearing a mask in public. While perceived mask effectiveness was a top predictor in the other country contexts, masks may have already been perceived as highly effective in Japan (27). Therefore, the top predictors of masking in Japan aligned with those observed for the other health behaviors—willingness to isolate and perceived severity. Altogether, the results indicate that people respond to health threats in a similar way across country contexts.

In addition to perceived severity and intervention effectiveness, demographic features such as gender and age had a significant effect on the uptake of health behaviors, with women and those of a higher age being more likely to practice the behavior, as has been found in other studies, but to a lesser extent than other key features (5,6,9,28). Similarly, trust and confidence in the government were significant predictors of the health behaviors in some contexts but were not top predictors.

Trust in the government and science has appeared as a determinant of health protective behavior in other research, but second to fear of disease (5,9,29,30). Recent research has linked COVID-19 health behavior uptake to the central components of the Theory of Planned Behavior— intentions, attitudes, perceived behavioral control, and subjective norms—as well as perceived risk and demographic features (28,31–34). While our study highlights perceived risk and touches on both attitudes (perceived intervention effectiveness) and behavioral control (ability to wear a mask), subjective norms are largely absent. Other research suggests a link between social network size and social distancing behavior, wherein greater exposure to subjective norms could influence behavior change (35,36).

Ryan et al. used similar machine learning models to understand predictors of mask-wearing behavior in Australia using the same Imperial College London YouGov behavior survey data (37). In agreement with our analysis, Ryan et al. identified willingness to isolate, perceived threat, and age as major determinants of masking, as well as survey week number—which is likely correlated with COVID-19 epidemic levels in the country. Similarly, Taye et al. used a random forest machine learning algorithm on a different behavioral dataset, combined with COVID-19 epidemiological metrics and the OxCGRT stringency index, across five European countries, and found COVID-19 cases, deaths, and the stringency index were the greatest predictors of self-protective behaviors during the pandemic (38). Urmi et al. using another COVID-19 behavioral dataset in the United States, found correlations between self-protective behaviors—like avoiding contact with others— and COVID-19 mortality, providing further evidence on the feedback loop between epidemic progression and adaptive human behavior (21).

This analysis has several limitations to consider. First, our analysis transformed health behavior measures into binary categories, which may fail to capture the nuance of human behavior as it exists on a continuum. Second, the behavioral survey data was cross-sectional and can only indicate population level changes in behavior, but not individual level changes. Third, observations that contained any missing variables were omitted instead of using imputation methods, and the possibility of bias was not considered. Fourth, the number of observations and the study period in our analysis was further restricted by when key questions were included in or omitted from the survey. Fifth, machine learning algorithms like XGBoost can be prone to overfitting, which is why we used Bayesian optimization and cross-validation for optimal hyperparameter and boosting round selection. Finally, machine learning algorithms cannot identify true causality with their predictor selection but given the alignment between these results and the literature, as well as the nature of the relationships between these variables, causation is plausible.

Parameterizing the feedback loop between epidemic spread and health behavior adoption has been a consistent challenge in transmission modelling. Gozzi et al. highlights two mechanistic approaches to this problem, data-driven feedback models and analytical driven feedback models (14). The former requires empirical data, which makes them more useful for hindsight, but not future projections, while the latter have remained theoretical without proper parameterization or validation. Our approach seeks to merge these two mechanisms. In future work, we will train the XGBoost algorithm on a simpler set of variables from real-world data, to be integrated directly into a network-based transmission model. The algorithm will be used to predict individual agent behavior based on personal attributes and epidemic progression. Such a model could then be used to explore how to increase health behavior uptake in the population through communicatory interventions.

## Data Availability

Instructions for accessing the publicly available data and the code for analysis are available at https://github.com/jmchev/covid-hb-determinants-machine-learning-analysis.

## Ethics

The data analyzed in this study are publicly available (16). The survey was conducted independently by the Imperial College London Big Data Analytical Unit in collaboration with YouGov Plc, neither of which has any affiliation with the authors or the present study. The current research constitutes a secondary analysis of de-identified and anonymized, publicly accessible data; therefore, institutional review board approval and informed consent were not required in accordance with applicable regulations and institutional policies.

## Author Contribution

Conceptualization (JMC, BJ, MH, RC, NDM, MK). Methodology (JMC, FvD, BJ, MH, RC, NDM, MK). Data Curation (JMC). Investigation (JMC, LS, LS, SW, FvD). Formal Analysis (JMC, MK). Validation (JMC, MK). Visualization (JMC, MK). Writing—original draft (JMC). Writing—review and editing (JMC, LS, LS, SW, FvD, LK, NL, BJ, US, ACV, MH, RC, NDM, MK). Supervision (MK). Funding acquisition (MK). Project administration (MK).

## Acknowledgements & Funding

This research was supported by a grant from ZonMW (The Netherlands Organisation for Health Research and Development). Project number: 10710062310022.

## References

1. Responding to community spread of COVID-19 [Internet]. [cited 2025 Dec 9]. Available from: https://www.who.int/publications/i/item/responding-to-community-spread-of-covid-19

2. Alyafei A, Easton-Carr R. The Health Belief Model of Behavior Change. In: StatPearls [Internet]. Treasure Island (FL): StatPearls Publishing; 2025 [cited 2025 Feb 7]. Available from: http://www.ncbi.nlm.nih.gov/books/NBK606120/ PubMed PMID: 39163427.

3. Ajzen I. The theory of planned behavior. Organ Behav Hum Decis Process. 1991 Dec 1;Theories of Cognitive Self-Regulation 50(2):179–211. doi:10.1016/0749-5978(91)90020-T

4. Balla J, Hagger MS. Protection motivation theory and health behaviour: conceptual review, discussion of limitations, and recommendations for best practice and future research. Health Psychol Rev. 2025 Jan 2;19(1):145–71. doi:10.1080/17437199.2024.2413011 PubMed PMID: 39420632.

5. Bish A, Michie S. Demographic and attitudinal determinants of protective behaviours during a pandemic: a review. Br J Health Psychol. 2010 Nov;15(Pt 4):797–824. doi:10.1348/135910710X485826 PubMed PMID: 20109274; PubMed Central PMCID: PMC7185452.

6. Hanratty J, Bradley DT, Miller SJ, Dempster M. Determinants of health behaviours intended to prevent spread of respiratory pathogens that have pandemic potential: A rapid review. Acta Psychol (Amst). 2021 Oct;220:103423. doi:10.1016/j.actpsy.2021.103423 PubMed PMID: 34624664; PubMed Central PMCID: PMC8492069.

7. Hanratty J, Leonard R, O’Connor SR, Keenan C, Chi Y, Ferguson J, et al. Psychological and psychosocial determinants of COVID related distancing behaviours: A systematic review. Campbell Syst Rev. 2024;20(4):e1442. doi:10.1002/cl2.1442

8. Leonard R, O’Connor SR, Hanratty J, Keenan C, Chi Y, Ferguson J, et al. Psychological and psychosocial determinants of COVID related face covering behaviours: A systematic review. Campbell Syst Rev. 2024;20(3):e1422. doi:10.1002/cl2.1422

9. Cabrera-Álvarez P, Hornsey MJ, Lobera J. Determinants of self-reported adherence to COVID-19 regulations in Spain: social norms, trust and risk perception. Health Promot Int. 2022 Dec 1;37(6):daac138. doi:10.1093/heapro/daac138 PubMed PMID: 36300700; PubMed Central PMCID: PMC9620366.

10. Funk S, Gilad E, Watkins C, Jansen VAA. The spread of awareness and its impact on epidemic outbreaks. Proc Natl Acad Sci U S A. 2009 Apr 21;106(16):6872–7. doi:10.1073/pnas.0810762106 PubMed PMID: 19332788; PubMed Central PMCID: PMC2672559.

11. Funk S, Gilad E, Jansen VAA. Endemic disease, awareness, and local behavioural response. J Theor Biol. 2010 May 21;264(2):501–9. doi:10.1016/j.jtbi.2010.02.032

12. Perra N, Balcan D, Gonçalves B, Vespignani A. Towards a Characterization of Behavior-Disease Models. PLOS ONE. 2011 Aug 3;6(8):e23084. doi:10.1371/journal.pone.0023084

13. Ryan M, Brindal E, Roberts M, Hickson RI. A behaviour and disease transmission model: incorporating the Health Belief Model for human behaviour into a simple transmission model. J R Soc Interface. 2024 Jun 5;21(215):20240038. doi:10.1098/rsif.2024.0038

14. Gozzi N, Perra N, Vespignani A. Comparative evaluation of behavioral epidemic models using COVID-19 data. Proc Natl Acad Sci. 2025 Jun 17;122(24):e2421993122. doi:10.1073/pnas.2421993122

15. Alford J. Imperial News [Internet]. 2020 [cited 2025 Sep 19]. Open data hub launches to track global responses to COVID-19 | Imperial News | Imperial College London. Available from: https://www.imperial.ac.uk/news/196793/open-data-launches-track-global-responses/

16. YouGov-Data/covid-19-tracker [Internet]. YouGov Data; 2025 [cited 2025 Jul 23]. Available from: https://github.com/YouGov-Data/covid-19-tracker

17. Mathieu E, Ritchie H, Rodés-Guirao L, Appel C, Gavrilov D, Giattino C, et al. COVID-19 Pandemic. Our World Data [Internet]. 2020 Mar 7 [cited 2025 Oct 17]. Available from: https://ourworldindata.org/coronavirus

18. RIVM. COVID-19 dataset [Internet]. [cited 2025 Oct 15]. Available from: https://data.rivm.nl/covid-19/

19. Hale T, Angrist N, Goldszmidt R, Kira B, Petherick A, Phillips T, et al. A global panel database of pandemic policies (Oxford COVID-19 Government Response Tracker). Nat Hum Behav. 2021 Apr;5(4):529–38. doi:10.1038/s41562-021-01079-8

20. Lundberg SM, Lee SI. A unified approach to interpreting model predictions. In: Proceedings of the 31st International Conference on Neural Information Processing Systems. Red Hook, NY, USA: Curran Associates Inc.; 2017. p. 4768–77. (NIPS’17).

21. Urmi T, Pant B, Dewey G, Quintana-Mathe A, Lang I, Druckman J, et al. Characterizing population-level changes in human behavior during the COVID-19 pandemic in the United States. Proc Natl Acad Sci. 2025 Sep 16;122(37):e2500655122. doi:10.1073/pnas.2500655122

22. McColl K, Mueller J, Martin-Lapoirie D, Verger P, Heyerdahl LW, Ventelou B, et al. Understanding the interplay between epidemiological and social cognitive drivers of behaviour change during the Covid-19 pandemic. Sci Rep. 2025 Aug 20;15(1):30556. doi:10.1038/s41598-025-14644-2

23. Teslya A, Nunner H, Buskens V, Kretzschmar ME. The effect of competition between health opinions on epidemic dynamics. PNAS Nexus. 2022 Nov;1(5):pgac260. doi:10.1093/pnasnexus/pgac260 PubMed PMID: 36712334; PubMed Central PMCID: PMC9802282.

24. Noguchi S, Ishimaru T, Yatera K, Fujino Y, Zaitsu M, Tabuchi T. Factors influencing mask-wearing behavior in the context of COVID-19 severity risks in the post-COVID-19 era: a Japanese Nationwide Epidemiological Survey in 2023. Environ Health Prev Med. 30:41. doi:10.1265/ehpm.24-00138 PubMed PMID: 40436798; PubMed Central PMCID: PMC12127079.

25. Suzuki R, Iizuka Y, Sugawara H, Lefor AK. Wearing masks is easy but taking them off is difficult – A situation in Japan during COVID-19 pandemic and after. Dialogues Health. 2024 Feb 3;4:100172. doi:10.1016/j.dialog.2024.100172 PubMed PMID: 38516216; PubMed Central PMCID: PMC10953900.

26. Wada K, Oka-Ezoe K, Smith DR. Wearing face masks in public during the influenza season may reflect other positive hygiene practices in Japan. BMC Public Health. 2012 Dec 10;12:1065. doi:10.1186/1471-2458-12-1065 PubMed PMID: 23227885; PubMed Central PMCID: PMC3536629.

27. Morishima M, Kishida K. Understanding attitudes toward hygiene mask use in Japanese daily life by using a repeated cross-sectional survey. Work Read Mass. 2018;61(2):303–11. doi:10.3233/WOR-182801 PubMed PMID: 30373980.

28. Wollast R, Schmitz M, Bigot A, Brisbois M, Luminet O. Predicting health behaviors during the COVID-19 pandemic: A longitudinal study. PLOS ONE. 2024 Mar 15;19(3):e0299868. doi:10.1371/journal.pone.0299868

29. Bronfman N, Repetto P, Cisternas P, Castañeda J, Cordón P. Government Trust and Motivational Factors on Health Protective Behaviors to Prevent COVID-19 Among Young Adults. Int J Public Health. 2022;67:1604290. doi:10.3389/ijph.2022.1604290 PubMed PMID: 35496944; PubMed Central PMCID: PMC9045398.

30. Han Q, Zheng B, Cristea M, Agostini M, Bélanger JJ, Gützkow B, et al. Trust in government regarding COVID-19 and its associations with preventive health behaviour and prosocial behaviour during the pandemic: a cross-sectional and longitudinal study. Psychol Med. 2023 Jan;53(1):149–59. doi:10.1017/S0033291721001306 PubMed PMID: 33769242; PubMed Central PMCID: PMC8144822.

31. Wollast R, Schmitz M, Bigot A, Luminet O. The Theory of Planned Behavior during the COVID-19 pandemic: A comparison of health behaviors between Belgian and French residents. PLOS ONE. 2021 Nov 4;16(11):e0258320. doi:10.1371/journal.pone.0258320

32. Bigot A, Banse E, Cordonnier A, Luminet O. Sociodemographic, Cognitive, and Emotional Determinants of Two Health Behaviors during SARS-CoV-2 Outbreak: An Online Study among French-Speaking Belgian Responders during the Spring Lockdown | Psychologica Belgica [Internet]. 2021 Jan 19. doi:10.5334/pb.712

33. Anaki D, Sergay J. Predicting health behavior in response to the coronavirus disease (COVID-19): Worldwide survey results from early March 2020. PLOS ONE. 2021 Jan 7;16(1):e0244534. doi:10.1371/journal.pone.0244534

34. Kojan L, Burbach L, Ziefle M, Calero Valdez A. Perceptions of behaviour efficacy, not perceptions of threat, are drivers of COVID-19 protective behaviour in Germany. Humanit Soc Sci Commun. 2022 Mar 24;9(1):97. doi:10.1057/s41599-022-01098-4

35. Marroquín B, Vine V, Morgan R. Mental health during the COVID-19 pandemic: Effects of stay-at-home policies, social distancing behavior, and social resources. Psychiatry Res. 2020 Nov;293:113419. doi:10.1016/j.psychres.2020.113419 PubMed PMID: 32861098; PubMed Central PMCID: PMC7439968.

36. Liu W, Sidhu A, Beacom AM, Valente TW. Social Network Theory. In: The International Encyclopedia of Media Effects [Internet]. John Wiley & Sons, Ltd; 2017 [cited 2025 Sep 24]. p. 1–12. Available from: https://onlinelibrary.wiley.com/doi/abs/10.1002/9781118783764.wbieme0092 doi:10.1002/9781118783764.wbieme0092

37. Ryan M, Ye J, Sexton J, Hickson RI, Brindal E. Face mask mandates alter major determinants of adherence to protective health behaviours in Australia. R Soc Open Sci. 2025 Mar 26;12(3):241941. doi:10.1098/rsos.241941

38. Taye AD, Borga LG, Greiff S, Vögele C, D’Ambrosio C. A machine learning approach to predict self-protecting behaviors during the early wave of the COVID-19 pandemic. Sci Rep. 2023 Apr 14;13(1):6121. doi:10.1038/s41598-023-33033-1

