## Appendix for "Identifying the determinants of health protective behaviors during the COVID-19 pandemic using machine learning: an analysis of six countries"

### Supplementary Appendix

**Table S1**. Total number of observations and demographic characteristics throughout the survey weeks (24 June 2020 to 13 January 2021) in each country following the removal of participants with missing observations.

| **Country** | **N** | **Age** *(Mean ± SD)* | **Gender** *% Female* | **Survey Weeks** |
|---|---|---|---|---|
| Canada | 10,029 | 50 ± 17 | 55 | 13 |
| France | 10,796 | 49 ± 16 | 53 | 13 |
| Japan | 4,345 | 48 ± 15 | 48 | 12 |
| Netherlands | 5,611 | 49 ± 17 | 53 | 12 |
| Sweden | 11,723 | 49 ± 17 | 51 | 13 |
| United Kingdom | 15,283 | 49 ± 17 | 52 | 17 |

**Figure S1.** Number of COVID-19 hospital patients per 100,000 population by country during the survey period from June 2020 to January 2021.

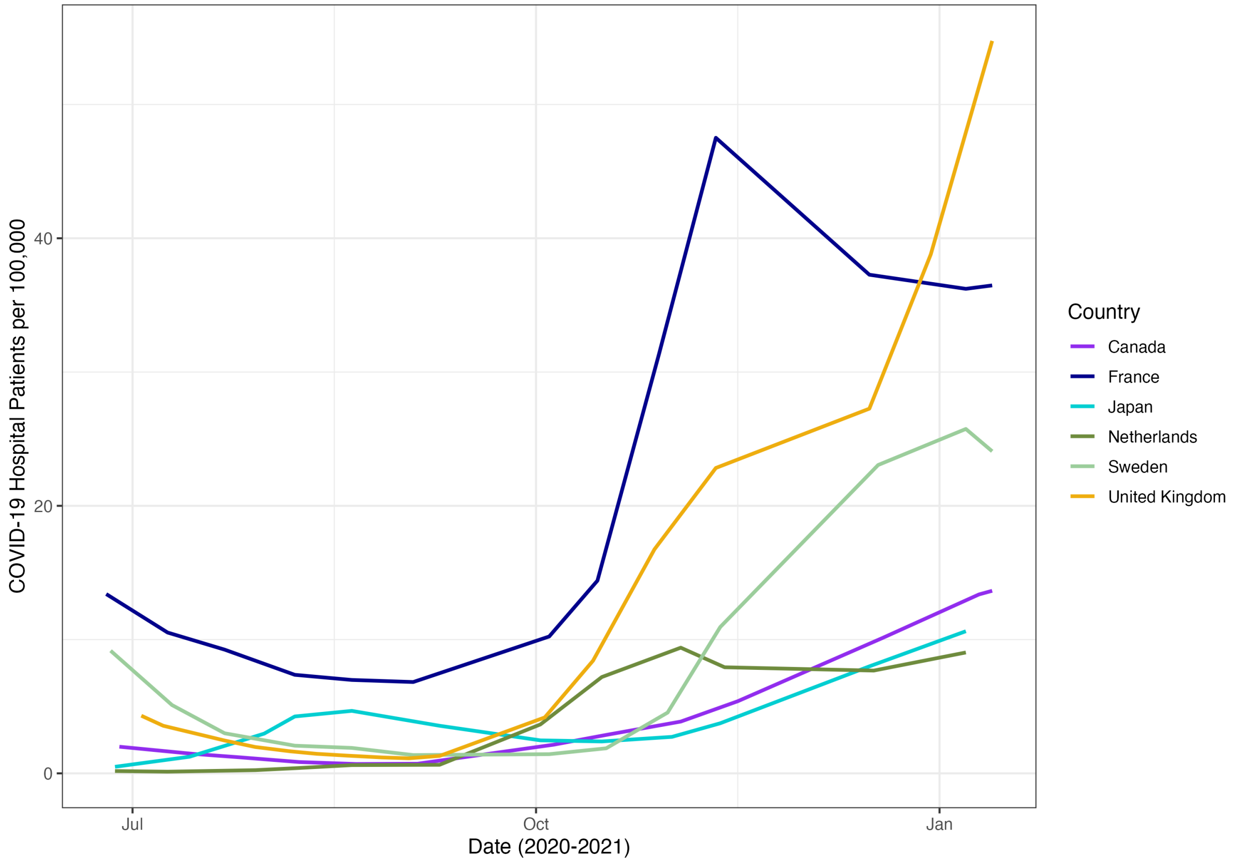

**Figure S2.** Average self-reported composite social distancing score (out of 20; blue) and the national stringency index (salmon) by country during the survey period from June 2020 to January 2021.

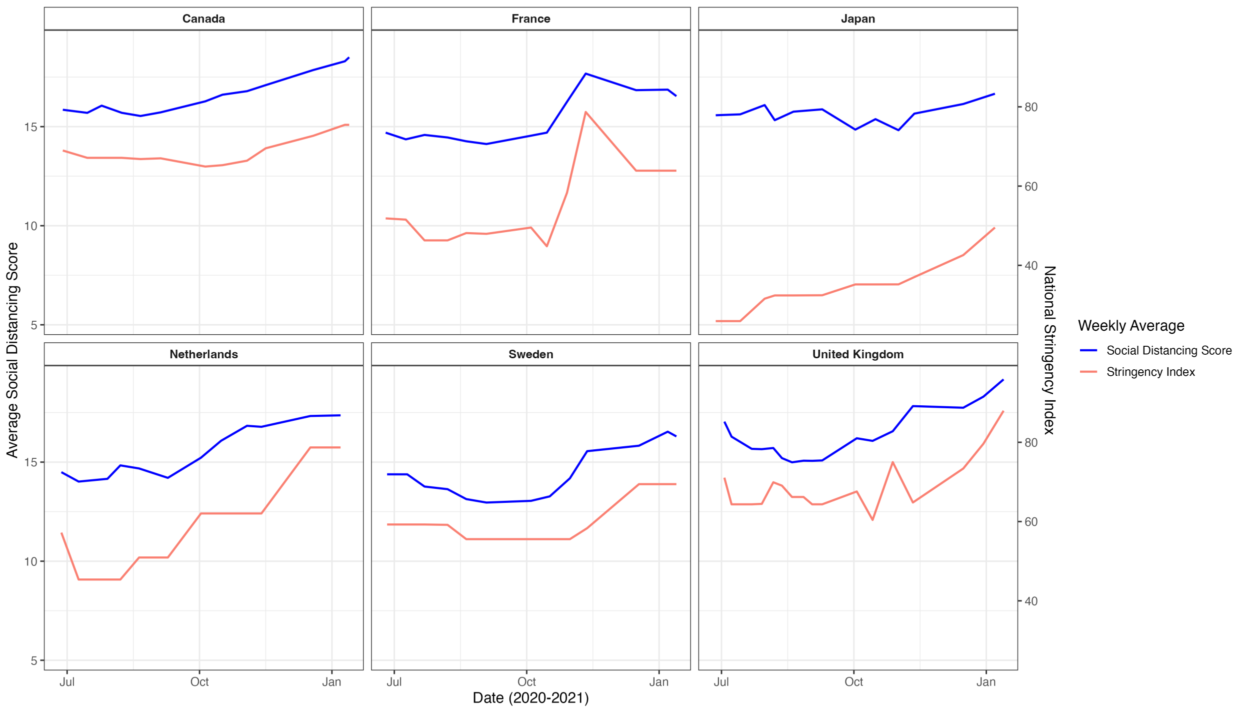


**Figure S3**. Average self-reported masking in public score on a 5-point Likert scale (blue) and the national stringency index (salmon) by country during the survey period from June 2020 to January 2021.

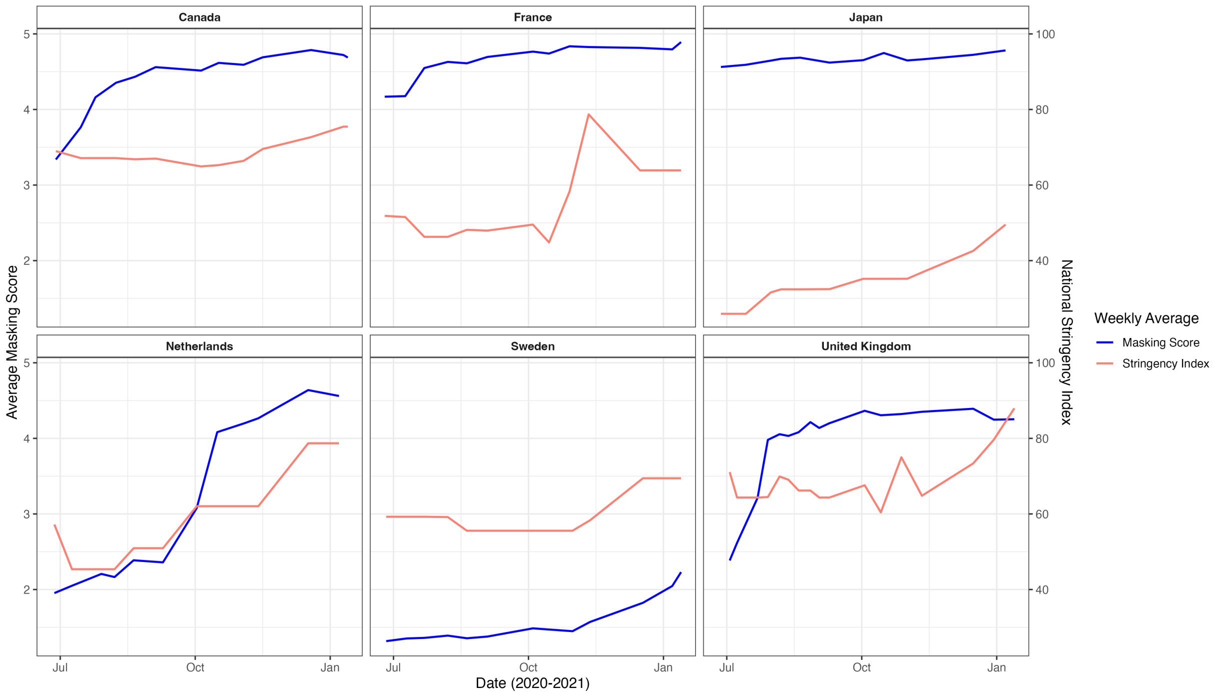

**Figure S4.** Average self-reported composite personal protective measure (PPM) score (out of 20; blue) and the national stringency index (salmon) by country during the survey period from June 2020 to January 2021.

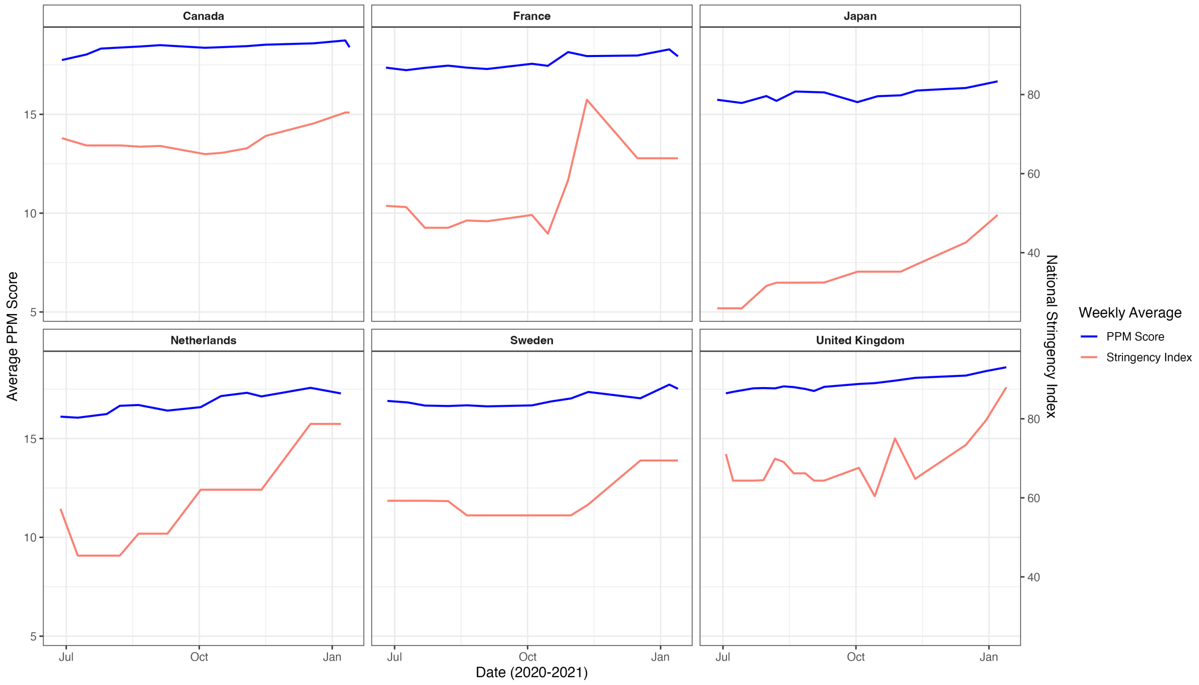


**Figure S5**. Binary distribution (proportion practicing) of self-reported health behaviors by country during the analysis period. A score of 16 was the binary cutoff for social distancing and personal protective measure (PPM) behaviors and a score of 4 was the binary cutoff for masking behavior.

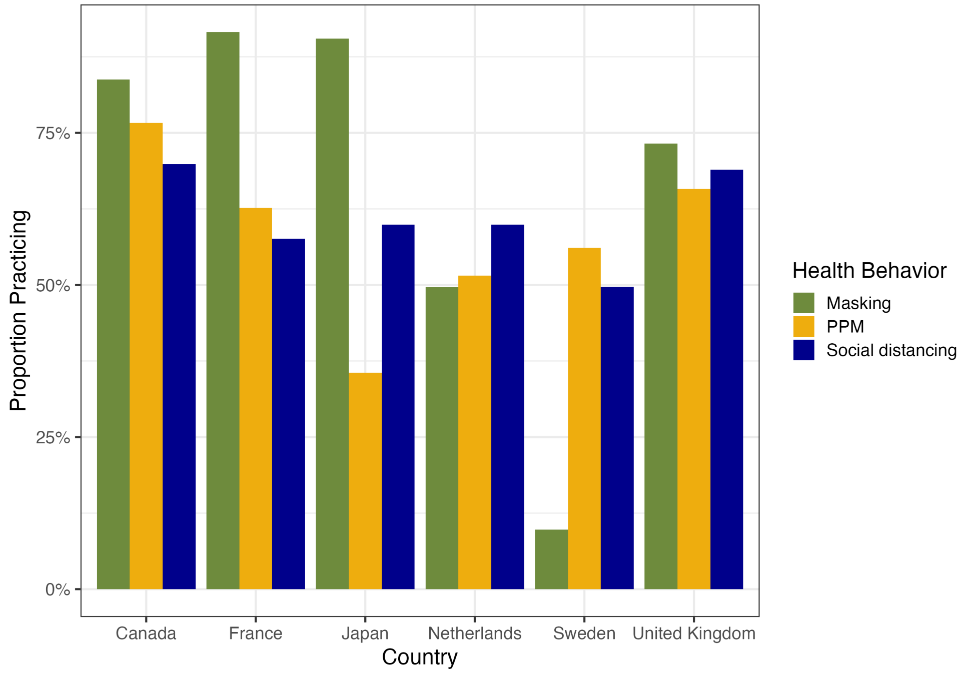

**Figure S6.** Inverse linear relationship between average composite social distancing score and average number of self-reported contacts from outside the home, after removing outliers (contacts > 100), stratified by working outside of the home, by country.

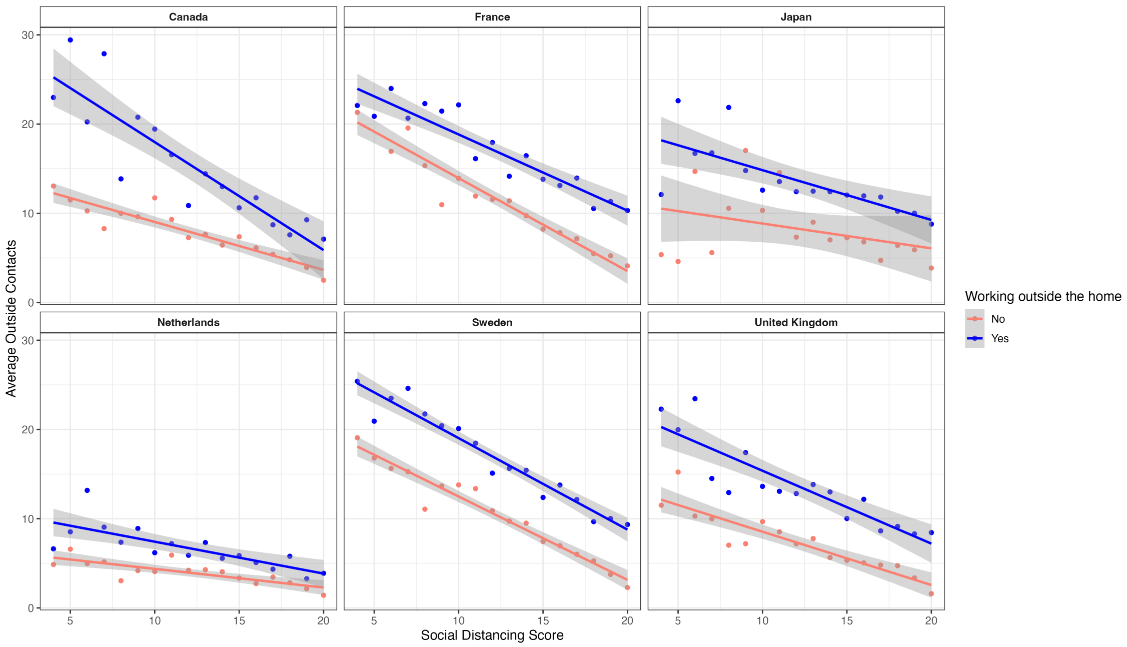


**Figure S7**. Standardized odds ratios and 95% confidence intervals for features that were significant predictors of social distancing behavior in logistic regression with stepwise AIC (p>0.05), when the national stringency index is included and COVID-19 hospital patients per 100,000 are excluded, by country.

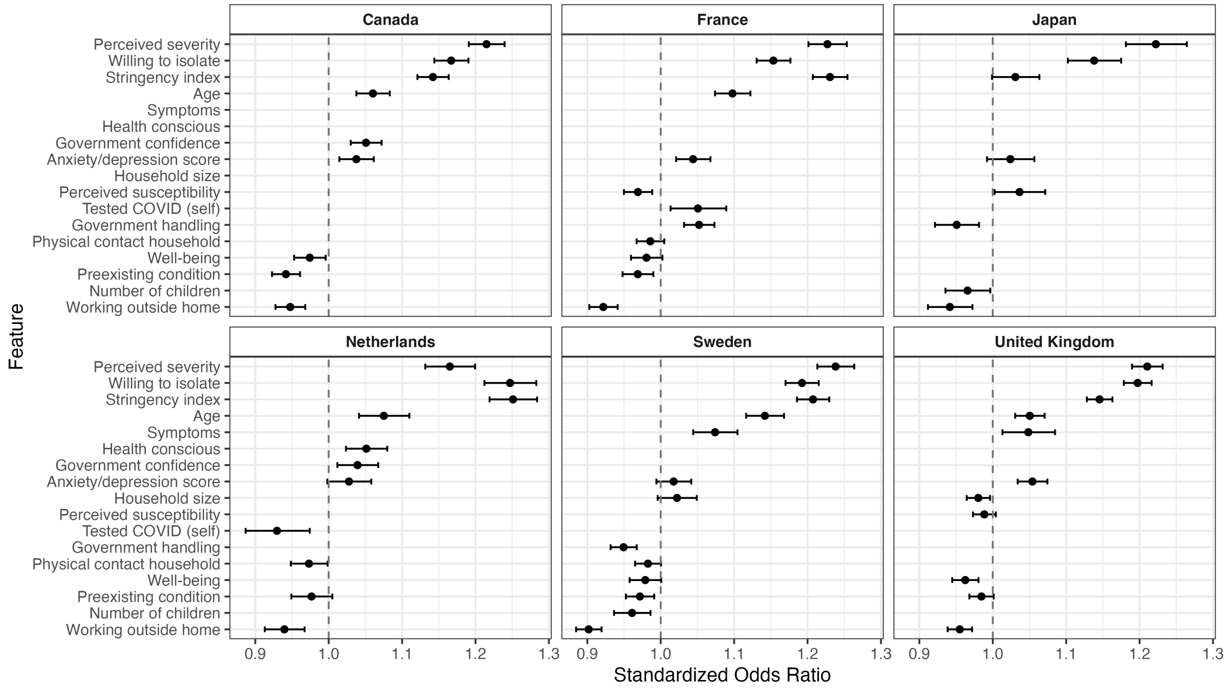

**Figure S8**. Average SHAP feature interaction values for social distancing by country.

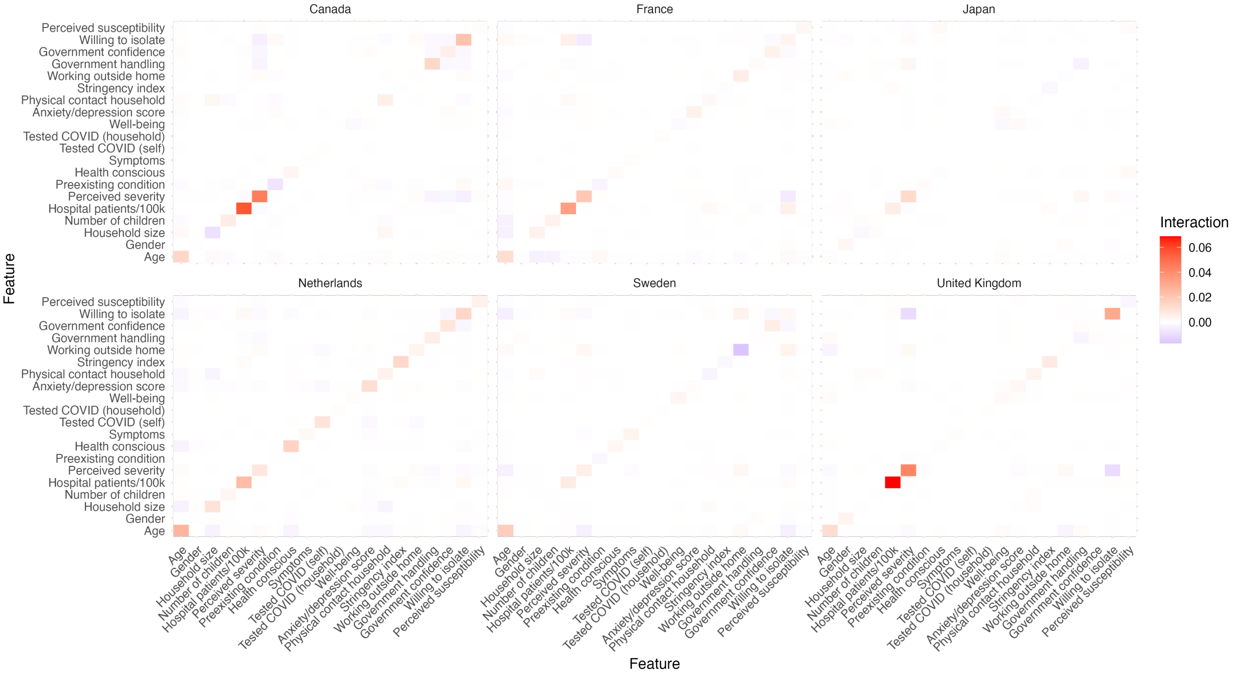


**Figure S9**. Standardized odds ratios and 95% confidence intervals of features that were significant predictors of masking behavior in logistic regression with stepwise AIC (p>0.05).

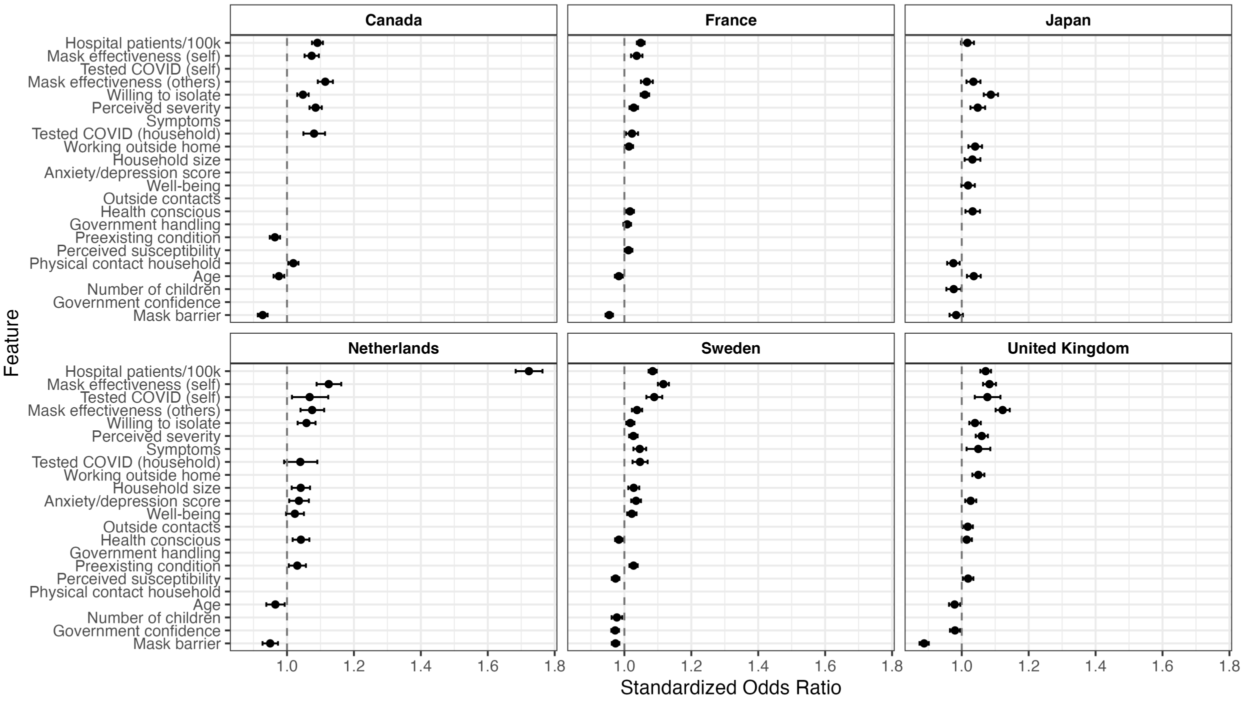

**Figure S10**. Standardized odds ratios and 95% confidence intervals of features that were significant predictors of increased PPM behavior in logistic regression with stepwise AIC (p>0.05).

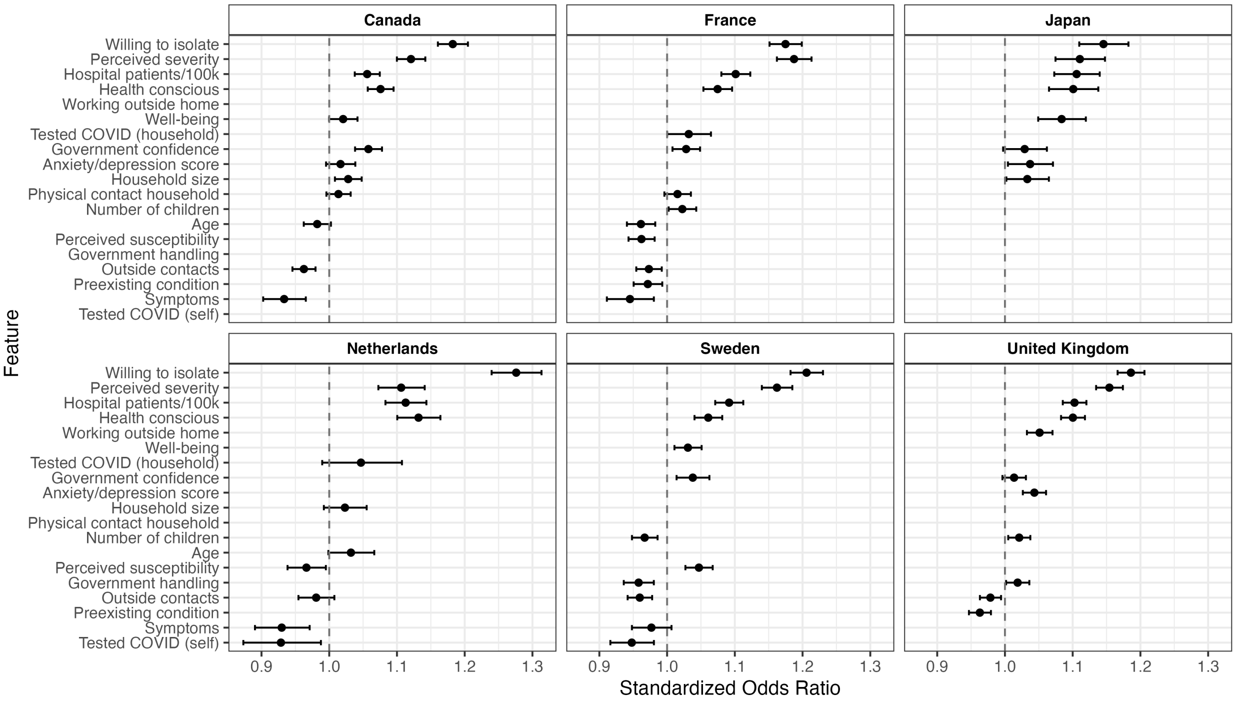
